# Discovery and Cross-Omic Replication of a Candidate Plasma Proteomic Signature for Anti-TNF Response in Crohn’s Disease

**DOI:** 10.64898/2026.11.18.26368439

**Authors:** Sudhir Ghandikota, Anil G. Jegga, Rebekah Karns, Jeffrey S. Hyams, Brendan Boyle, Joshua Noe, Kimberly Jackson, Kimberly Lynch, Jonathan R. Dillmann, Alex Nasr, Thomas Aviles, Lee A. Denson, Phillip Minar

## Abstract

**Background and Aims:** Despite expanding therapeutic options for Crohn’s disease (CD), treatment selection remains largely empiric. Identifying biomarkers linking inflammatory endotypes to treatment outcomes could inform first-line therapy selection. We sought to identify baseline plasma proteins associated with clinical, biochemical, and endoscopic outcomes in patients with CD receiving anti-TNF therapy.

**Methods:** In this prospective multicenter cohort, children and young adults with CD initiating infliximab or adalimumab underwent plasma proteomic profiling at baseline and months 3 and 12 using SomaScan v4.1 (7,322 analytes) with deep remission (clinical and endoscopic) as the primary outcome. Rank-based composite correlation analysis identified candidate proteins associated with prespecified treatment outcomes, followed by functional enrichment analyses. Regularized machine learning models were used to derive and evaluate a candidate protein signature.

**Results:** In a cohort of 80 participants, mean age 13.7 (SD 3.5) years, 42.5% female, and 90% White, composite rank analysis identified 73 candidate proteins associated with one or more outcomes. Seven candidate proteins (*IFNA6, IGHM, MBL2, UBASH3A, REG1A, GABBR2,* and *NTS*) were retained for evaluation. In an independent adult cohort (n=324), models based on whole-blood expression of the seven corresponding genes achieved mean AUPRCs of 0.71 (baseline) and 0.76 (week 14) for the cohort-defined primary response outcome.

**Conclusions:** Plasma proteomic profiling identified a candidate 7 protein signature associated with anti-TNF treatment outcomes in CD. Cross-omic replication demonstrated consistent outcome-associated signals in an independent cohort. Although orthogonal validation of the protein signature is needed, these findings support the potential of proteomic biomarkers to guide biologically informed treatment selection.

## Introduction

Crohn’s disease (CD) is a chronic, relapsing inflammatory disorder resulting from dysregulated interactions among host genetics, the immune system, and environmental exposures.^1^ Although a cure remains elusive, advances in biologic and small molecule therapeutics have substantially improved outcomes by targeting key inflammatory pathways. For more than two decades, inhibitors of tumor necrosis factor-α (anti-TNF), including infliximab and adalimumab, have been a cornerstone of treatment for moderate-to-severe pediatric CD and are the most commonly prescribed first-line biologic therapy.

The early use of anti-TNF therapy, coupled with strategies to optimize drug exposure, has reduced CD-related surgery and improved rates of sustained steroid-free remission.^2–4^ In real-world practice, while more than 75% of patients achieve an initial clinical response, complete intestinal healing occurs in fewer than 40%, and 10-15% lose response annually despite dose optimization.^5, 6^ Importantly, currently available biomarkers cannot prospectively identify patients whose underlying inflammatory biology is unlikely to respond to long-term TNFα inhibition, leaving clinicians to determine treatment failure only after adequate drug exposure has been achieved. Consequently, patients with pharmacodynamic failure may spend months undergoing dose optimization before switching to an alternative advanced therapy with a different mechanism of action. With the recent approval of ustekinumab for pediatric CD, the expanding availability of biologics targeting distinct inflammatory pathways further underscores the need for predictive biomarkers that can identify patients most likely to benefit from specific therapeutic mechanisms. The urgency of this approach is further reinforced by evidence that treatment response declines with each successive advanced therapy, while ongoing bowel inflammation can lead to irreversible fibrotic remodeling. Together, these challenges highlight a critical need for biomarkers or companion diagnostics to guide earlier, mechanism-based treatment selection within the expanding therapeutic armamentarium.^7^

Although first-line biologic selection is often informed by clinical phenotype, disease location, severity, and complications, these features incompletely capture the underlying inflammatory heterogeneity that may determine therapeutic response.^8, 9^ Emerging studies suggest that patients with CD exhibit distinct molecular endotypes that may enable more personalized treatment selection. Martin et al. identified a pathogenic cellular module associated with anti-TNF nonresponse using single-cell RNA sequencing,^10^ while Ungaro et al. identified serum protein signatures associated with disease progression in newly diagnosed children with CD.^11^ Defining the patient-specific endotypes before treatment could enable selection of therapies whose mechanisms of action align with the dominant inflammatory pathways driving bowel inflammation in individual patients.

Advances in high-throughput plasma proteomics now enable comprehensive characterization of circulating proteins and provide a promising platform for biomarker discovery.^12^ Unlike earlier studies limited by small cohorts or targeted protein panels, these technologies allow unbiased profiling of thousands of circulating proteins from a small volume of peripheral blood, offering an opportunity to identify clinically relevant biomarkers of therapeutic response.^12–17^

In this study, we used the SomaScan platform to quantify more than 7,300 plasma proteins from 80 children and young adults with CD initiating infliximab or adalimumab. Our primary aim was to identify baseline plasma proteins associated with month-12 (M12) deep remission and characterize enriched biological pathways. Using rank-based composite correlation and machine learning-based approaches, we identified a 7-protein signature and evaluated its association with multiple treatment outcomes.

## Materials and Methods

### Cohort

The Precision Crohn’s Disease Management Utilizing Predictive Protein Panels (ENvISION) study (NCT04131504) is a multicenter, prospective observational study of children and young adults with CD initiating anti-TNF biologic therapy (infliximab or adalimumab). Participants entered the longitudinal ENvISION study through either the diagnostic or treatment enrollment pathway. In the diagnostic pathway, participants were enrolled before their initial ileocolonoscopy and followed to establish a final diagnosis of CD, ulcerative colitis, or a non-inflammatory bowel disease (IBD) condition. Participants subsequently diagnosed with CD who initiated anti-TNF therapy were then eligible for the longitudinal ENvISION cohort study. In the treatment pathway, participants with an established diagnosis of CD were enrolled immediately before initiating anti-TNF therapy and entered the longitudinal ENvISION cohort directly. All participants in the longitudinal cohort were anti-TNF naïve, younger than 19 years of age, and followed prospectively for one year. Baseline demographic characteristics, disease phenotype and severity, and prior corticosteroid or immunomodulator exposure were recorded.

Participants underwent longitudinal collection of research-only blood and stool biospecimens throughout the first year of therapy. Follow-up ileocolonoscopy was performed after one year of anti-TNF therapy as part of routine clinical care at each participating center. If a clinically-indicated follow-up ileocolonoscopy was not performed, participants were offered a research-only colonoscopy. An optional research-only magnetic resonance enterography (MRE) was also offered to all participants one year from treatment initiation.

For both biologics, the initial dose selection and maintenance intervals were at the discretion of the treating physician at their respective medical center. Dose or interval optimizations occurred throughout maintenance and were largely based on clinical assessments and a combination of proactive or reactive TDM to maintain a trough concentration >5 µg/mL for both biologics.^18^

Patients were enrolled at Cincinnati Children’s Hospital Medical Center, Connecticut Children’s Hospital, Nationwide Children’s Hospital, and at the Medical College of Wisconsin between October 2019 and December 2022. All subjects or their guardians provided informed consent for this study before collecting any study data. The study was approved by the Institutional Review Board at all four participating centers.

Healthy controls (HC) were recruited at Cincinnati Children’s Hospital Medical Center and frequency-matched to the CD cohort by age and sex. HC were 6-19 years of age and were required to have no chronic medical conditions, no antibiotic exposure within the preceding four weeks, fecal calprotectin <50 µg/g, and no chronic gastrointestinal symptoms meeting Rome IV criteria for a functional gastrointestinal disorder.^19^

### Primary Outcome

Participants were evaluated prospectively for up to one year or until discontinuation of the index anti-TNF biologic or CD-related surgery, whichever occurred first. At M12, clinical, biochemical, radiologic, and endoscopic outcomes were assessed. The primary outcome was M12 deep remission, defined as the combination of clinical and endoscopic remission while remaining on the same anti-TNF therapy without surgery. Clinical remission is defined as a weighted pediatric CD activity index (wPCDAI) <12.5 with endoscopic remission defined as a Simple Endoscopic Score-Crohn’s Disease (SES-CD) <3.^20^ The SES-CD was independently scored by three blinded reviewers (AN, TA, PM), each of whom reviewed the complete, de-identified ileocolonoscopy video without knowledge of prior SES-CD scores or treatment outcomes. Inter-rater agreement was assessed and scoring discrepancies were resolved through repeat blind review by all three reviewers.

### Secondary Outcomes: Early Response and Remission

Early treatment outcomes were assessed at month-3 (M3). Primary clinical nonresponse was defined as no improvement or worsening from baseline wPCDAI or a M3 wPCDAI >40. Primary biochemical nonresponse was defined as a failure to achieve a reduction in fecal calprotectin of >100 µg/g among participants with baseline concentrations >250 µg/g or a reduction in C-reactive protein (CRP) of ≥0.5 mg/dL among those with baseline CRP >1.0 mg/dL. Steroid-free clinical remission was defined as a wPCDAI<12.5 without corticosteroid use. Fecal calprotectin remission was defined as a fecal calprotectin concentration <250 µg/g. Biochemical remission was defined as a fecal calprotectin <250 µg/g or, when unavailable, a CRP level <0.5 µg/dL. Adequate anti-TNF exposure was defined according to the American Gastroenterological Association Institute guidelines as an infliximab trough concentration ≥5 µg/mL at the first maintenance infusion (dose4) or an adalimumab trough concentration ≥7.5 µg/mL during maintenance.^21^

Exploratory M3 outcomes of interest included fecal calprotectin healing (<150 µg/g) and ideal anti-TNF exposure, defined as an infliximab trough concentration ≥4.7 µg/mL or an adalimumab trough concentration ≥14 µg/mL, with drug concentration thresholds derived from cohort-specific exposure-response relationships for deep remission.^22^

### Secondary Outcomes: Late Remission

Late treatment outcomes were assessed at M12. Steroid-free clinical remission was defined as a wPCDAI <12.5 without corticosteroid use for >8 weeks. Endoscopic remission was defined by a SES-CD <3. Deep remission was also assessed among those who remained on the index anti-TNF therapy and underwent repeat endoscopy, excluding those who discontinued therapy or underwent surgery. Additional M12 outcomes included fecal calprotectin remission, combined fecal calprotectin and endoscopic remission, and combined clinical, fecal calprotectin and endoscopic remission as defined above. For the combined remission outcomes, participants who discontinued anti-TNF therapy or underwent CD-related surgery before M12 were considered not in remission. Transmural remission was assessed using the simplified Magnetic Resonance Index of Activity (sMARIA), scored by a single experienced radiologist (JRD), and defined as a sMARIA<2.^23^ Adequate anti-TNF exposure was defined as a trough concentration ≥5 µg/mL for infliximab and ≥7.5 µg/mL for adalimumab.^21^

Exploratory M12 outcomes included fecal calprotectin healing (<150 µg/g), transmural healing, and ideal anti-TNF exposure. Participants who discontinued anti-TNF therapy or underwent CD-related surgery before M12 were classified not achieving transmural healing, regardless of MRE availability. Ideal anti-TNF exposure at M12 was defined as an infliximab trough concentration ≥4.7 µg/mL or an adalimumab trough concentration ≥14 µg/mL, based on cohort-specific exposure-response relationships for deep remission.^22^

### Peripheral Blood Proteomics

Peripheral blood was collected longitudinally using P100 blood collection tubes (BD Biosciences, Franklin Lakes, NJ) to stabilize plasma proteins and stored at -80° Celsius for batched analyses. Plasma proteomics was performed by the Washington University School of Medicine Genome Technology Access Center (St. Lous, MO) using the SomaScan® v4.1 platform (SomaLogic, Inc., Boulder, CO). SomaScan employs Slow Off-rate Modified Aptamers (SOMAmers) to measure 7,322 proteins. Protein abundance was semi-quantified as relative fluorescence units (RFU), generated by DNA hybridization microarrays following SOMAmer binding to target proteins. Standard normalization, scaling, calibration, and quality control procedures were applied to minimize technical variability. The assay provides a broad, reproducible dynamic range spanning femtomolar to micromolar protein concentrations.^24^

### Additional Biomarker Investigations

Fecal calprotectin was measured at baseline, M3, and M12 using a commercial ELISA (Buhlmann, Switzerland).^25^ CRP values were obtained from routine clinical laboratory testing and TDM for both biologics was assessed at M3 and M12 using a drug-tolerant electrochemiluminescence immunoassay (Esoterix, LabCorp specialty lab, Calabasas, CA).

### Statistical Analysis

#### Composite Correlation Analysis

To identify candidate proteins associated with each outcome, Pearson correlation coefficients and corresponding asymptotic Student’s *t*-test p-values were calculated between protein abundance and binary outcome variables.^26^ Proteins were ranked within each outcome by two-sided P-value. A composite rank was calculated as the harmonic mean of the outcome-specific ranks. Outcomes included in the composite were weighted equally. The top 1% of proteins by composite rank were designated exploratory candidates and evaluated using functional enrichment analysis. Functional enrichment analysis was performed using ToppFun application in the ToppGene Suite^27^. Enrichment P-values were adjusted using the Benjamini and Hochberg method. Networks linking candidate proteins to enriched functional terms were visualized in Cytoscape.^28^

### Regularized regression analysis

Regularized logistic regression models were developed using penalized maximum likelihood estimation to evaluate the predictive ability of candidate proteins.^29^ Specifically, generalized linear models (GLMs) incorporating an elastic net penalty were fit across a grid of regularization parameter values. Protein abundances were used to predict each study outcome. Additionally, we implemented a leave-one-out cross-validation (LOOCV) approach to identify the best parameter values for the trained models. In each training experiment, samples were randomly divided into training and test sets. The training sets were for model fitting and parameter tuning, whereas the test set was reserved for model evaluation.

Model performance was evaluated using the area under the precision-recall curve (AUPRC) and the area under the receiver operating characteristic curve (AUROC). Individual GLMs were then developed for each candidate protein. To obtain robust performance estimates, 10 randomized training and test splits were performed, and performance metrics were averaged across all experiments. All GLMs were adjusted for baseline treatment exposures, disease phenotype, and treatment-related variables. We used a similar framework for our validation experiments, training models on 70% of samples and evaluating on the remaining 30%, with LOOCV to select the best parameter values. We trained 100 random models to report robust validation performance metrics.

### Cross-omic Verification

The PANTS study is a multicenter, prospective observational cohort of 1,610 anti-TNF naïve patients with CD initiating infliximab or adalimumab in the United Kingdom. Participants (aged ≥6 years) and followed through 54 weeks, with clinical, biochemical, and treatment outcomes assessed at baseline and weeks 14, 30, and 54.^30^ In a post-hoc analysis, investigators performed whole-blood RNA-seq in a subset of 324 PANTS participants who met the following inclusion critiera, age ≥16 years, and a baseline CRP ≥4 mg/L and/or baseline fecal calprotectin >100 µg/g.^31^ Primary response was assessed at week 14 and defined as a decrease in CRP to ≤3 mg/L or by ≥50% from baseline, and a decrease in the Harvey-Bradshaw index to ≤4 points or by ≥3 points from baseline.^31^ Furthermore, the primary response subjects were required to be in remission (CRP of ≤3 mg/L and HBI score of ≤ 4 points, no ongoing steroid therapy, and no exit due to treatment failure) at weeks 30 and 54.^31^

### Ethical Considerations

All subjects and/or their guardians provided informed consent for this study before collecting any study data. The study was approved by the Institutional Review Board at all four participating centers.

## Results

### Patient cohort

We enrolled 111 participants through the diagnostic enrollment pathway, of whom 64 were diagnosed with CD. Among these, 42/64 initiated anti-TNF therapy and consented to the longitudinal ENvISION study. Separately, 38 participants with established CD were enrolled through the treatment enrollment pathway immediately before initiating infliximab or adalimumab, as their first biologic therapy. Together, these two pathways yielded a final longitudinal cohort of 80 participants, including 63 who initiated infliximab and 17 who initiated adalimumab (see Supplementary Figure 1). Of the 80 participants, 34 (42.5%) were female, 72 (90%) were White, and the mean (SD) age at anti-TNF initiation was 13.7 (3.5) years. Before anti-TNF initiation, 37 participants (46.3%) were receiving corticosteroids, including 19 receiving prednisone and 18 receiving oral budesonide. Most participants (90%) initiated anti-TNF therapy without a concomitant immunomodulator. The HC group comprised the first 30 consecutively enrolled participants who met eligibility criteria.

At M12 follow-up, 69 participants (86.3%) remained on the same anti-TNF biologic, with five (6.3%) undergoing CD-related surgery and six (7.5%) discontinuing the index anti-TNF therapy. Among participants who remained on the same biologic without surgery, 59/69 (85.5%) underwent a repeat ileocolonoscopy, while 44/80 (55%) underwent the research-only MRE. Baseline characteristics were generally balanced between the infliximab and adalimumab treatment groups, although participants treated with infliximab were younger and had a lower body mass index at study entry, as previously published.^22^

#### Primary Outcome

Among the 70 participants included in the primary outcome analysis, 48.6% achieved deep remission, with participants who discontinued anti-TNF therapy or underwent CD-related surgery classified as not achieving deep remission. Deep remission was comparable between participants receiving infliximab (50.9%) and adalimumab (38.5%, p=0.54). Baseline characteristics were generally similar between participants with and without deep remission (Table 1). Those who achieved deep remission had a higher starting adalimumab dose (mg/kg), lower wPCDAI, and a longer interval from CD diagnosis and anti-TNF initiation.

**Table 1.**
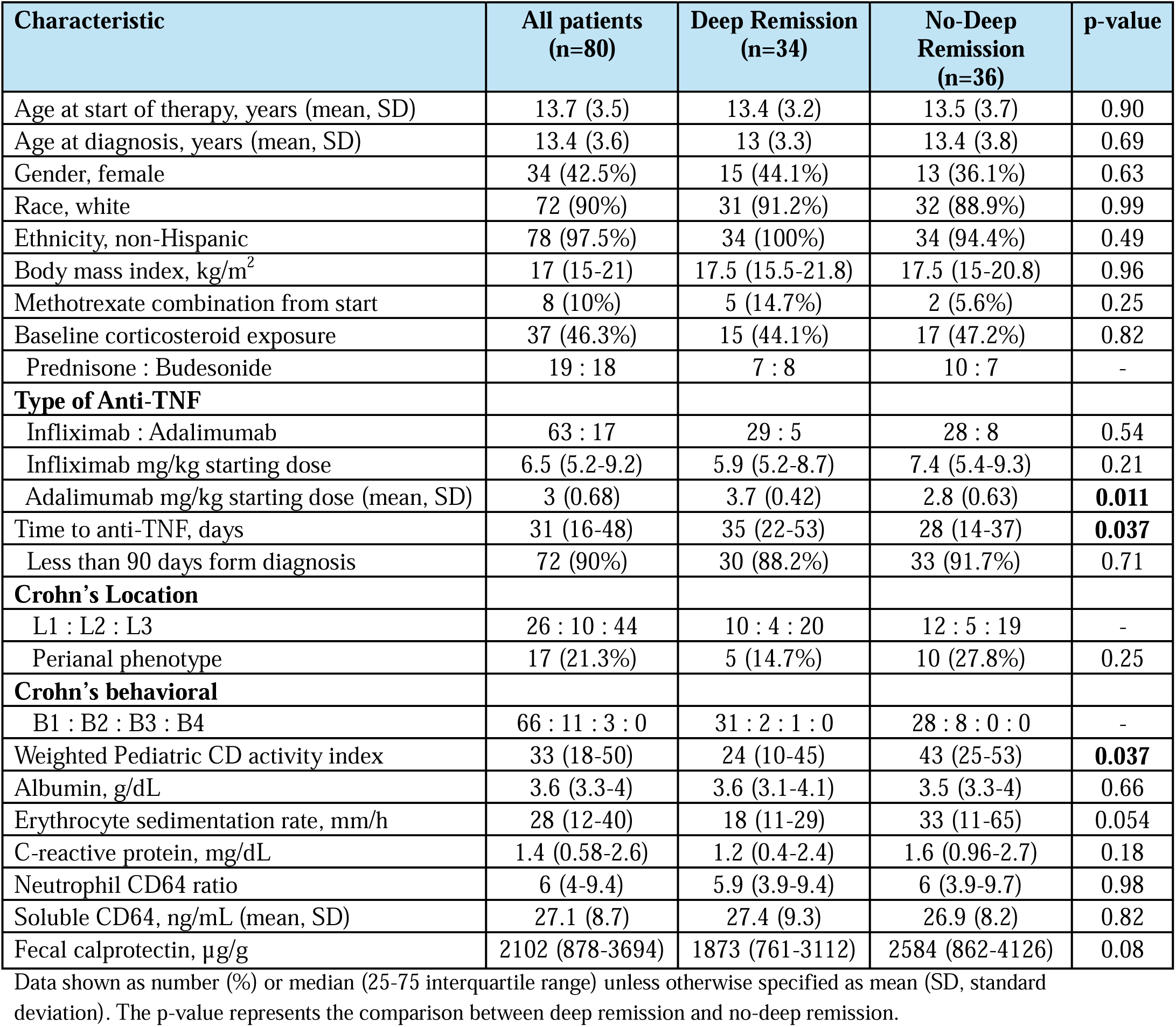
Baseline demographics, disease characteristics, and laboratory results for all subjects and those who achieved deep remission and those who did not achieve deep remission.

#### Secondary Outcomes

Clinical, biochemical, endoscopic, and transmural outcomes at M3 and M12 were similar between participants treated with infliximab and adalimumab (see Supplementary Table 1). At M12, 73.8% of participants in the ENvISION cohort achieved steroid-free clinical remission, 53.1% achieved fecal calprotectin remission, 54.5% achieved transmural remission, and 39.3% achieved combined clinical, fecal calprotectin and endoscopic remission (see Supplementary Figure 2). Outcomes were also similar between participants receiving anti-TNF monotherapy and combination therapy (data not shown).

### Protein Discovery: Data preprocessing and Normalization

Protein abundance measurements were used to conduct a composite correlation analysis to identify candidate proteins associated with different M3 and M12 outcomes (see Supplementary Table 1) following anti-TNF administration. Before analysis, we retained proteins meeting the prespecified detection or quality criterion in at least 50% of samples at each sampling time (Figure 1). We applied variance stabilization normalization (VSN) to reduce the influence of technical noise in the final protein measurements.^32^ VSN applies a parametric transformation intended to reduce the dependence of variance on mean signal (see Supplementary Figure 3).^33^

**Figure 1:**
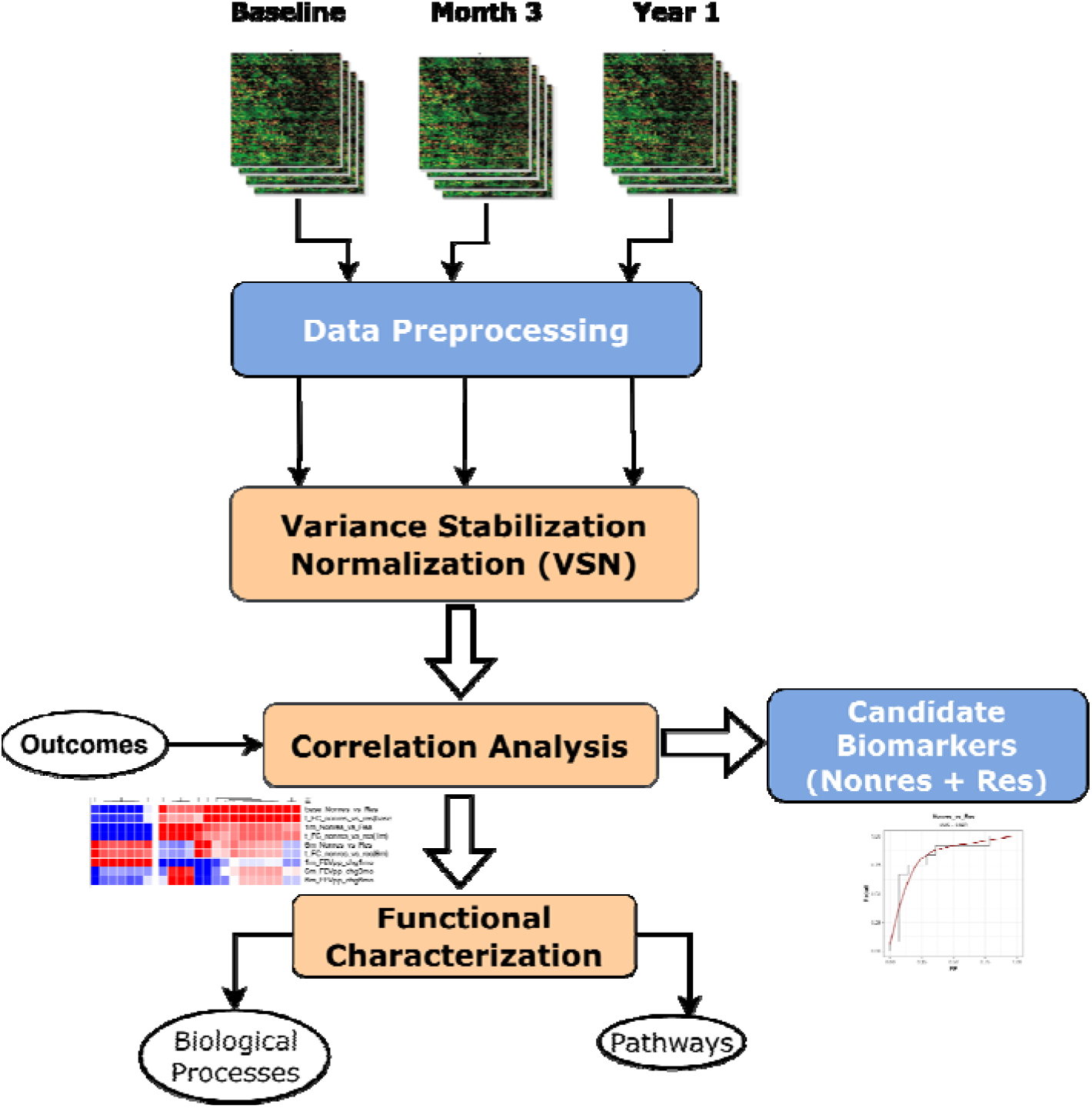
Study and analysis workflow: Protein measurements from three different time points are processed by implementing variance stabilization normalization to remove the influence of technical noise. Correlation analysis is then applied to the normalized data to identify significant candidate proteins and potential biomarkers predictive of the anti-TNF outcomes at baseline. Functional enrichment analysis is also applied to identify the biological processes and pathways associated with the correlated proteins.

### Correlated Proteins

The VSN-normalized data from each time point were used in the composite correlation analysis. The analysis included twenty-one distinct outcomes assessed at M3 and M12 following anti-TNF initiation, encompassing clinical, biochemical, endoscopic and transmural measures (see Supplementary Table 1). Student’s asymptotic p-values were used to assess the significance of the correlation scores. Based on the composite ranks derived from these p-values (see Materials and Methods), we identified 73 candidate proteins (top 1%) that were strongly correlated with multiple outcomes (Figure 2, see Supplementary File 1). Of these 73 candidates, 43 showed a favorable direction of association with treatment outcomes, whereas 30 showed correlations in an unfavorable direction. Compared with HC, 49 candidate proteins were differentially expressed in CD participants (adjusted p-value <0.05) and 43 proteins were significantly downregulated in CD (Figure 2).

**Figure 2:**
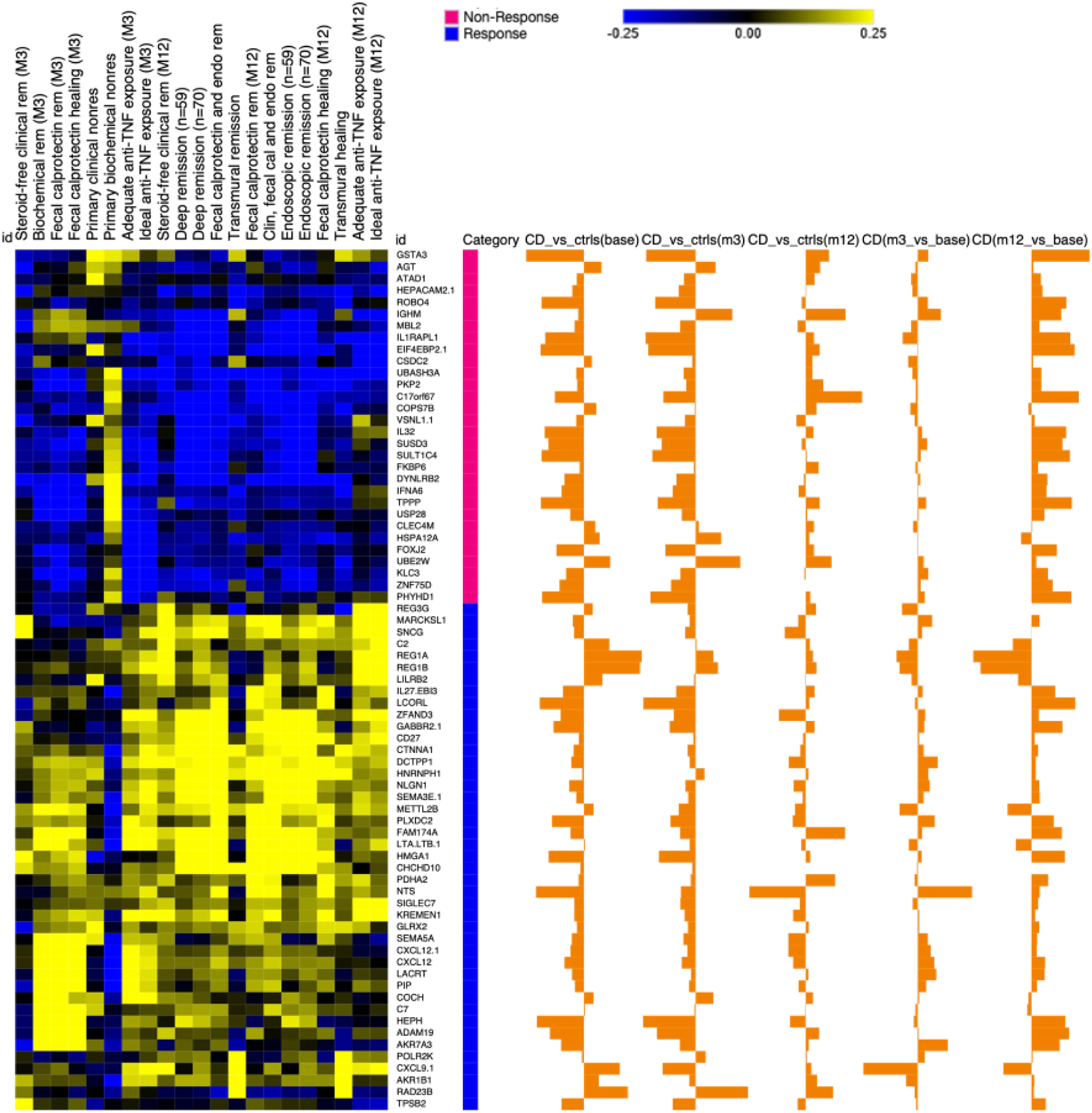
Outcome correlation heatmap: Correlation heatmap of the 73 correlated proteins (rows) based on the composite significance p-value-based ranks (top 1%). Yellow color represents positive correlations, while the blue color in the heatmap indicates negative correlations with the outcome variables (along the columns).

Functional enrichment analysis of the 73 candidate proteins identifed terms related to adaptive immune response (11 proteins; FDR-adjusted p-value =1.15E-02), humoral immune response (13 proteins; FDR-adjusted p-value =2.47E-07), cytokine activity (8 proteins; FDR-adjusted p-value =2.15E-04), cell-cell signaling (16 proteins; FDR-adjusted p-value =1.01E-02), and cell adhesion (17 proteins; FDR-adjusted p-value =7.27E-03, Figure 3). Additional enriched terms included IL6/JAK/STAT3 signaling (4 proteins; FDR-adjusted p-value =3.57E-02), complement activation (4 proteins; FDR-adjusted p-value =3.9E-03), regulation of endocytosis (7 proteins; FDR-adjusted p-value =2.08E-02), the matrisome (17 proteins; FDR-adjusted p-value =3.59E-06) and the extracellular matrix (8 proteins; FDR-adjusted p-value =1.61E-05, Figure 3, see Supplementary File 1).

**Figure 3:**
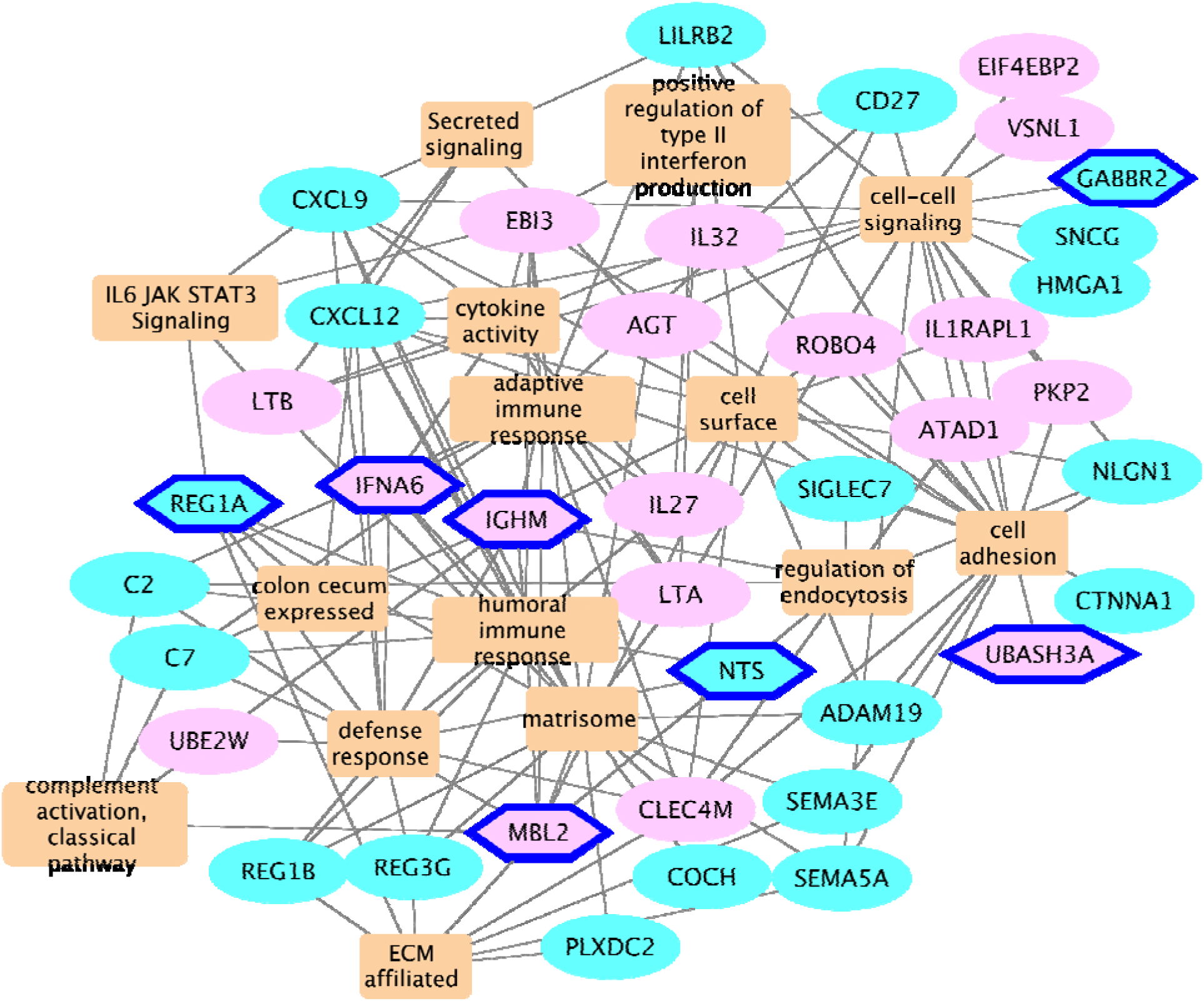
Functional enrichment network of the 73 candidate proteins: Circular and hexagon-shaped nodes represent proteins, and rectangular nodes denote enriched annotation terms meeting an FDR-adjusted p-value <0.05. Proteins correlated with anti-TNF response are highlighted using the pink colored nodes, while the sky-blue nodes represent the proteins associated with non-response. Proteins represented using navy-blue bordered hexagons are the 7-protein biomarkers.

### Biomarker candidates

To derive a candidate protein panel, we fit elastic net logistic regression models^34^ using baseline (pre-treatment) protein abundance to predict each M3 and M12 outcome (see Materials and Methods). We trained more than 1,400 penalized logistic regression models, adjusted for pretreatment exposures, type of anti-TNF biologic, and disease phenotype, and computed AUC scores for all protein-outcome pairs (Figure 4A). These scores reflect a protein’s ability to predict a specific anti-TNF outcome status before starting biologic therapy (see Supplementary File 1). Seven candidate proteins (*IFNA6, IGHM, MBL2, UBASH3A, REG1A, GABBR2, and NTS*) achieved a mean AUPRC of 0.75 across more than two outcomes. Four proteins (*IGHM, IFNA6, MBL2,* and *UBASH3A*) were associated with nonresponse, and three (*GABBR2, REG1A, and NTS*) were associated with response to anti-TNF (Figure 4B).

**Figure 4:**
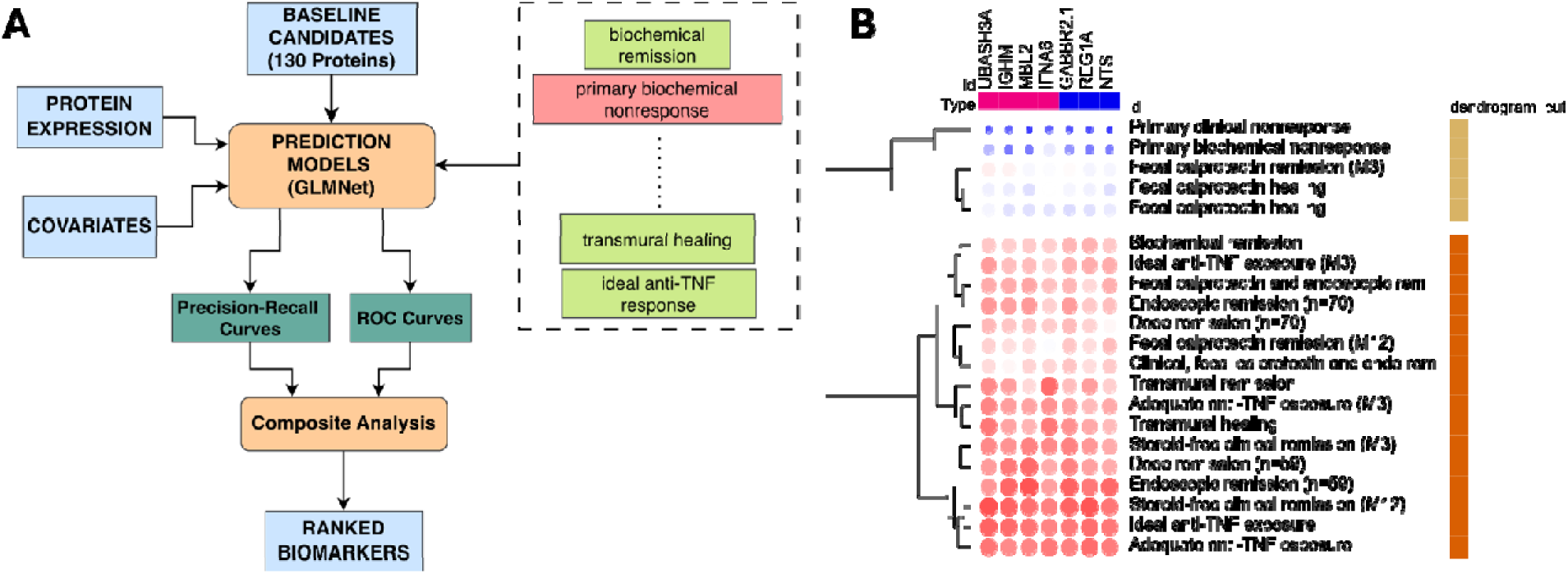
Regularized regression analysis and biomarker candidates. (A). Schematic representation of the workflow used for selecting biomarkers, where GLMs were trained for each protein-outcome pair. The AUC scores from each experiment were used to rank and filter the final protein biomarker list. (B). Heatmap of the AUPRC scores for the 7-protein biomarker panel across the 21 selected outcomes. The AUPRC scores are averaged over 10 different experimental runs.

The combined 7-biomarker model demonstrated predictive performance across multiple M12 outcomes, including steroid-free clinical remission (AUPRC = 0.91 ± 0.026; AUROC = 0.77 ± 0.056), combined fecal calprotectin and endoscopic remission (AUPRC = 0.78 ± 0.074; AUROC = 0.81 ± 0.068), deep remission (AUPRC = 0.8 ± 0.067; AUROC = 0.72 ± 0.075), and transmural remission (AUPRC = 0.79 ± 0.084; AUROC = 0.74 ± 0.082). The model demonstrated strong performance for those with ideal anti-TNF exposure concentrations (AUPRC = 0.88 ± 0.05; AUROC = 0.77 ± 0.078) at M12 (Figure 5). The model was also moderately predictive of M12 fecal calprotectin remission (AUPRC = 0.67 ± 0.06; AUROC = 0.63 ± 0.088). At M3, model performance was more modest for M3 steroid-free clinical remission (AUPRC = 0.75 ± 0.097; AUROC = 0.66 ± 0.099), fecal calprotectin remission (AUPRC = 0.82 ± 0.01; AUROC = 0.62 ± 0.14) and adequate anti-TNF exposure concentrations (AUPRC = 0.73 ± 0.098; AUROC = 0.63 ± 0.11, Figure 5).

**Figure 5:**
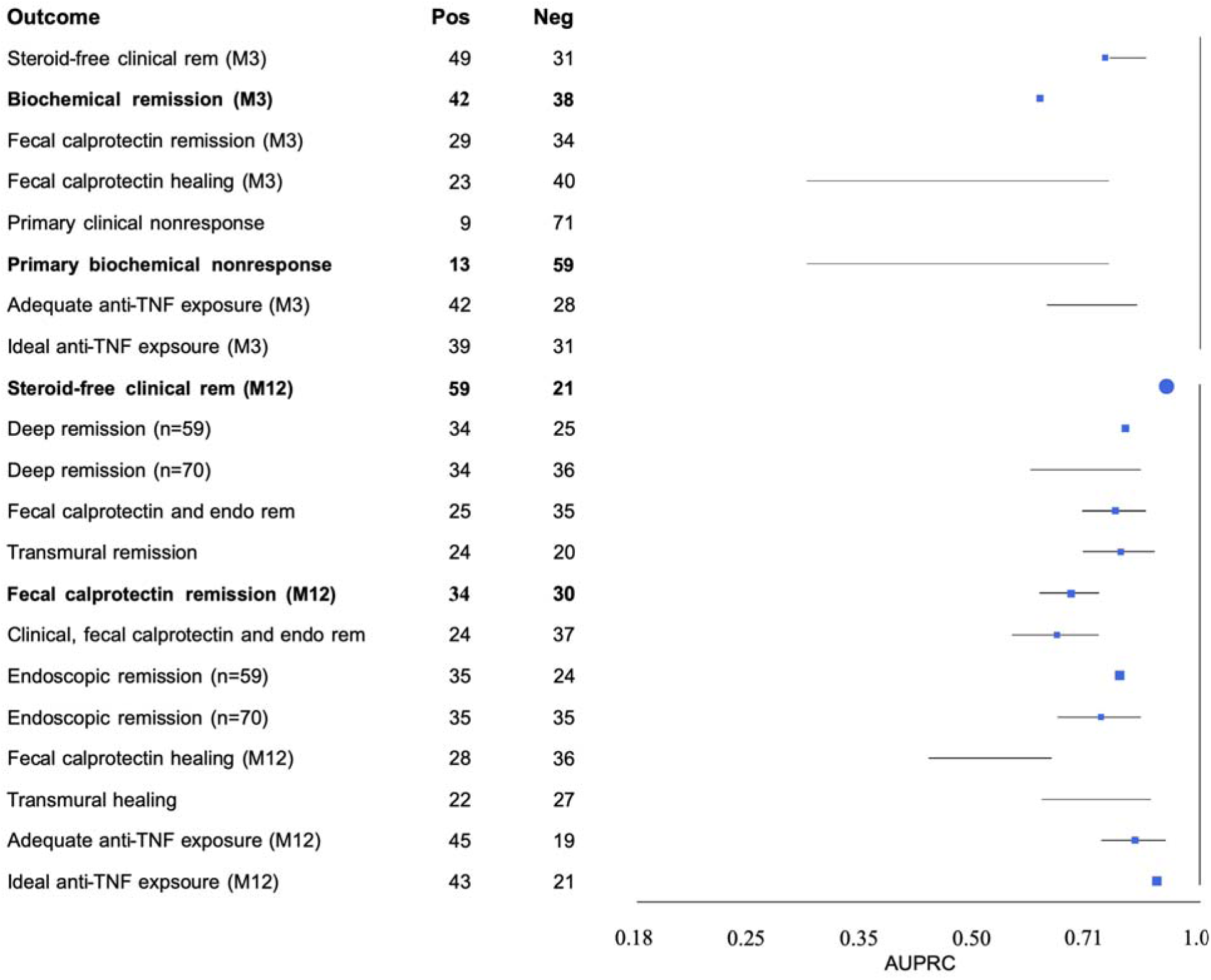
Forest plots of outcome predictions: AUPRC scores from models trained using the 7-protein biomarker panel in predicting each of the selected outcomes. The label (sample) counts associated with each outcome are also included in the forest plot. The outcomes most similar to the primary response from the PANTS cohort (cross-omic replication) are highlighted in bold.

### Cross-omic replication in an independent transcriptomic cohort

To determine whether gene expression corresponding to the seven candidate proteins was associated with anti-TNF response, we performed a cross-omic replication analysis using the independent PANTS prospective cohort,^31^ a prospective, UK-wide observational study of anti-TNF treatment outcomes. The post-hoc analysis included 814 whole blood RNA-sequencing samples from 324 patients collected across four sampling times. Raw RNA-seq counts were converted to counts per million (CPM) and log-transformed. Regularized logistic regression models incorporating the seven corresponding genes as covariates were trained to classify the cohort-defined primary response outcome (see Materials and Methods). Separate models were fitted at baseline and weeks 14, 30, and 54.

Model performance increased across successive study visits. At baseline, our trained biomarker model achieved an AUPRC score of 0.704 ± 0.044, which increased to 0.76 ± 0.07 at week 14 (corresponding to M3 in our cohort, Figure 6a-b). Performance remained high at weeks 30 and 54, with AUPRCs of 0.783 ± 0.073 and 0.818 ± 0.089, respectively (Figure 6c-d). These estimates were averaged across 100 independent experiments using distinct training and testing data splits (see Supplementary File 1). Models trained using PANTS data were adjusted for clinical and demographic characteristics, including age, sex, disease duration, smoking history, and body mass index.

**Figure 6:**
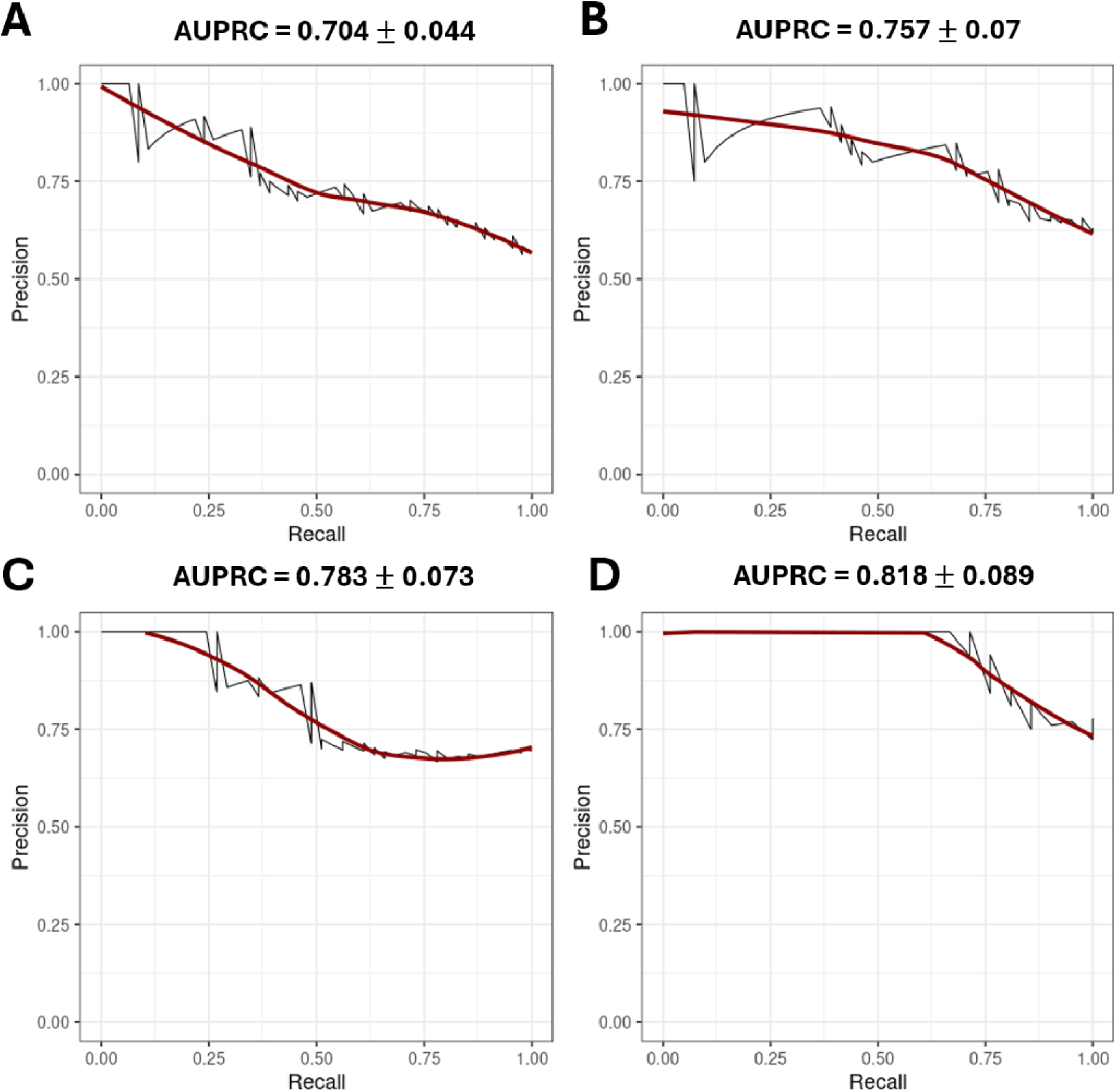
Cross-omic replication using blood transcriptomic data from the PANTS cohort: Precision-recall curves of the models trained on the blood gene expression data from an independent validation study (PANTS). The models were trained on data from four different time points: baseline (Panel A), week 14 (Panel B), week 30 (Panel C), and week 54 (Panel D). The AUPRC scores averaged over 100 different experimental runs are added as the plot labels.

## Discussion

With the rapid expansion of advanced therapies, identifying blood-based biomarkers that predict treatment response and enable pairing of patients to specific therapies remains a major research priority for advancing precision medicine in adult and pediatric IBD.^35–37^ Currently available blood and stool biomarkers, including CRP, albumin, and fecal calprotectin, provide important measures of inflammatory burden but do not directly identify the biological pathways driving an individual patient’s disease or reliably predict response to a specific therapeutic mechanism. This distinction may be particularly relevant for anti-TNF therapy, where primary or secondary nonresponse may reflect distinct mechanisms, including inadequate drug exposure due to accelerated clearance or immunogenicity (pharmacokinetic failure), persistent inflammation despite adequate drug exposure (pharmacodynamic failure), or inflammation driven by non-TNF-dependent pathways (mechanistic failure). Molecular profiling before and early after treatment, therefore, has the potential to complement conventional biomarkers by identifying biological features associated with subsequent treatment response, enabling earlier therapeutic optimization and potentially limiting progressive, irreversible bowel damage.^11, 38, 39^

In this study, unbiased plasma proteomic profiling identified 73 candidate proteins strongly correlated with distinct clinical, biochemical, endoscopic, radiologic, and pharmacologic outcomes associated with anti-TNF response or nonresponse. Of these proteins, 43 were associated with favorable treatment outcomes and 30 with unfavorable outcomes. Functional enrichment analysis identified immune, cytokine-signaling, complement, cell-adhesion, cell-cell signaling, and extracellular-matrix terms. These findings suggest that the proteomic correlates of anti-TNF response extend beyond generalized systemic inflammation and encompass several biological processes relevant to immune activation, tissue injury, and intestinal remodeling. Importantly, these associations are hypothesis-generating and do not establish that individual proteins or pathways causally determine anti-TNF response.

Complementary predictive modeling identified an exploratory 7-protein baseline panel that discriminated subsequent endoscopic remission and deep remission. The panel included proteins representing diverse biological processes, including immunoglobulin and complement-associated proteins (*IGHM* and *MBL2*), interferon signaling (IFNA6), antimicrobial and epithelial responses (REG1A), and T-cell signaling and regulation of cytokine production (UBASH3A), together with other proteins whose relationship to IBD treatment response has not been well characterized. The presence of proteins associated with both favorable and unfavorable outcomes within the same panel further supports the concept that anti-TNF response may reflect the balance of multiple biological processes rather than the abundance of a single inflammatory marker. Given the exploratory nature of these analyses, these findings require further validation in larger, more diverse cohorts, including orthogoinal validation using independent protein-level assays.

The 7-protein panel also discriminated M12 trough concentrations above versus below the prespecified thresholds. This finding is notable as drug exposure represents a distinct but closely related aspect of anti-TNF response. Anti-TNF concentrations are influenced by dosing, interval adjustment, patient adherence, as well as inflammatory burden and covariates of drug clearance such as albumin, body size (kg), and immunogenicity. Thus, some circulating proteins may identify biological features associated with pharmacokinetics and drug clearance, whereas other proteins may reflect pharmacodynamic response or persistent inflammatory activity despite adequate exposure.

As one of the first studies to utilize the 7,322-analyte SomaScan® proteomic platform in a large cohort of patients with CD, identifying an independent cohort with comparable protein-level data for validation was challenging. Therefore, the 7-protein panel was evaluated by assessing the expression of the corresponding genes in the independent PANTS cohort, providing cross-omic replication across a different patient population and molecular platform. The corresponding gene panel demonstrated increasing discrimination across study visits, with average AUPRC values of 0.704 at baseline, 0.76 at M3, 0.79 at week 30, and 0.82 at M12. However, replication at the transcript level does not constitute independent validation of the 7-protein signature. Independent protein-level validation, direct comparison with established clinical and biochemical predictors, and evaluation in cohorts receiving alternative advanced therapies with differing therapeutic mechanisms will be required to determine whether these biomarkers provide treatment-specific information and can be used to guide treatment selection.

Several proteins within the 7-protein panel have biological or previously reported associations with CD and intestinal inflammation, providing additional support for their investigation as candidate biomarkers. Deep sequencing of the B cell receptor (BCR) repertoire has identified enrichment of IGHM isotypes among CD-associated clones in blood and intestinal mucosa.^40^ Interestingly, this trend was not observed in HC or in patients with ulcerative colitis (UC). Mannose-binding lectin (MBL) protein and MBL-dependent complement activation have been implicated in inflammatory processes in CD.^41^ MBL2, a component of the lectin complement pathway, is also particularly relevant. MBL2 has been associated with intestinal inflammation in CD and, notably, was previously identified by intestinal tissue proteomics as being increased in patients with UC who did not respond to infliximab, providing an independent link between MBL2 and anti-TNF nonresponse.^42^ *GABBR2* represents another biologically plausible candidate, as GABAergic signaling regulates intestinal innate lymphoid cell activity and IL17A production, and disruption of this pathway exacerbates experimental colitis.^43^ Using data from over 34,000 IBD cases in a genome-wide association study stratified by sex, investigators identified *UBASH3A*, a T-cell-enriched immune regulatory gene, as a significant IBD-associated locus.^44^

Additional candidates may identify less well-characterized pathways relevant to treatment response. The regenerating family member 1 alpha (*REG1A*) gene is involved in the antimicrobial humoral immune response. Recent works have linked regenerating gene (REG) family genes to IBD-associated intestinal inflammatory response and epithelial regeneration.^45, 46^ Its role as a circulating marker of treatment response in CD remains to be established. *IFNA6* may implicate type I interferon signaling and adaptive immune response, whereas *NTS* functions as a neurotransmitter with prominent expression in the gastrointestinal tract. However, their roles in CD and anti-TNF response remain poorly defined. Mechanistic studies are needed to determine whether these proteins participate in treatment-response pathways or are correlates of baseline inflammation, disease severity, or drug disposition. Therefore, their identification within a multivariable panel should be considered hypothesis-generating, rather than evidence of a mechanistic role.

Our study has several strengths, including the prospective longitudinal design, broad plasma proteomic profiling, serial sampling following anti-TNF initiation, and comprehensive assessment of clinical, biochemical, pharmacokinetic, endoscopic, and radiologic outcomes. The inclusion of objective measures of intestinal inflammation and drug exposure allowed candidate proteins to be evaluated across multiple complementary dimensions of treatment response. Our analytical approach incorportate regularized machine learning models, repeated cross-validation, and evaluation of predicitve performance across multiple clinically relenat outcomes, providing a framework for identifying and priortizing candidate protein biomarkers. In addition, evaluation of the corresponding genes in an independent cohort of pediatric and adult patients with CD provided cross-omic evidence supporting the predictive utility of these biomarkers across age groups and molecular platforms.

Several limitations should also be considered. The cohort was modest relative to the number of proteins and outcomes examined, with smaller numbers of participants contributing to some outcome analyses. The overlap among the 21 outcomes and use of a top-1% candidate threshold may have amplified related biological signals. Participants who discontinued therapy, underwent surgery, or did not undergo M12 colonoscopy may have differed systematically from those with complete follow-up assessments. Treatment selection, dosing, interval adjustment, and therapeutic drug monitoring reflected clinical practice and were not standardized. The cohort was predominantly White, which limits generalizability of our findings to more diverse populations. Given the limited availability of independent assays for these seven candidate proteins, we did not undertake orthogonal validation of each protein. Finally, the independent PANTS analysis used whole-blood gene expression rather than direct plasma protein measurements and therefore represents cross-omic replication rather than independent protein-level validation of the candidate panel.

In conclusion, in a prospective pediatric and young-adult cohort initiating infliximab or adalimumab, broad plasma proteomic profiling identified a 7-protein candidate panel with promising predictive performance across multiple clinically relevant treatment outcomes. The corresponding genes demonstrated outcome-related discrimination in an independent adult and pediatric transcriptomic cohort, providing cross-omic support for the identified protein signature. Together, these findings highlight the potential of plasma proteomic profiling to identify molecular signatures predictive of anti-TNF treatment response before treatment initiation. Independent protein-level validation in larger, more diverse cohorts and subsequent evaluation across advanced therapies with differing mechanisms of action are essential next steps toward translating these findings into biomarkers that can guide individualized therapeutic selection and advance precision medicine for patients with CD.

## Data Availability

All data produced in the present work are contained in the manuscript and are also available upon reasonable request to the authors

## Acknowledgments

We sincerely thank the patients and their families for their participation and commitment throughout this one-year study. Their contributions, including completing study surveys and providing research-specific biospecimens, were essential to the success of this research.

## Ethical Considerations

Written informed consent was obtained from all participants or their legal guardians before any study-related data collection. The study was approved by the Institutional Review Board (2019-0730) at all four participating centers.

## Conference presentation

Portions of this manuscript were presented as an oral abstract presentation at the Digestive Disease Week, May 2024 in Washington, DC.

## Author contributions to manuscript

SG, AJ, and RK performed bioinformatics, statistical analysis, drafted and revised the manuscript. JSH, BB and JN contributed to study design, data collection, and revision of the manuscript. KJ contributed as project management, data acquisition and entry and revision of the manuscript. KL contributed as biospecimen collection, processing, analysis, and revision of the manuscript. JRD performed radiographic severity scoring and revision of manuscript, AN and TA performed outcome analysis and critical review of the manuscript. LAD contributed to study design and planning, data analysis, and critical revision of the manuscript. PM contributed to study concept and design, obtained funding, performed statistical analysis and interpretation, drafted and revised the manuscript.

## Data Transparency

Once published, the de-identified proteomic dataset will be made available to researchers upon request to the corresponding author.

