## Supplemental Data for "Discovery and Cross-Omic Replication of a Candidate Plasma Proteomic Signature for Anti-TNF Response in Crohn’s Disease"

**Supplemental Figure 1**. **ENvISION enrollment pathways**. Participants were enrolled either through the diagnostic pathway at the time of the ileocolonoscopy or through the treatment pathway immediately before initiation of anti-TNF therapy.

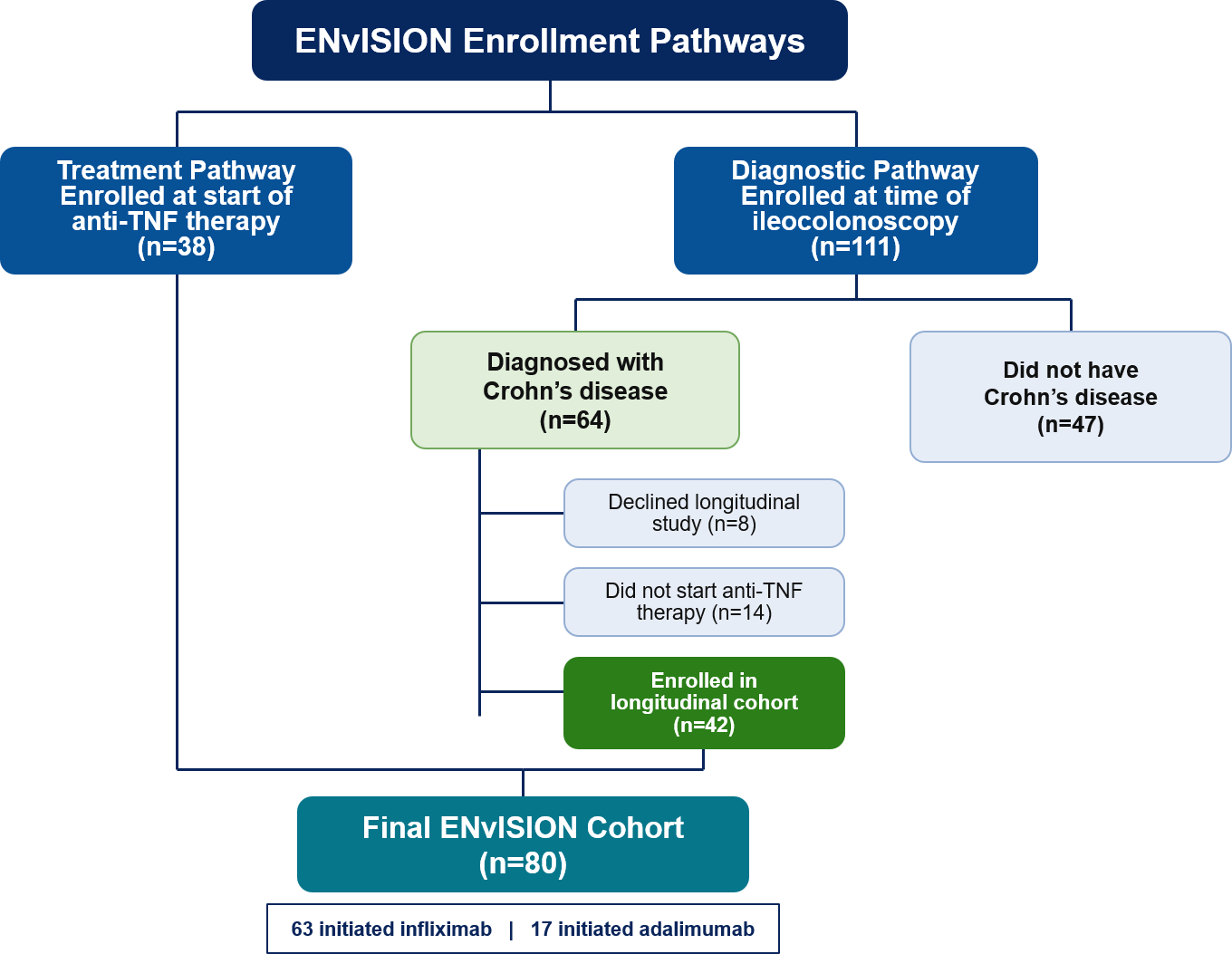

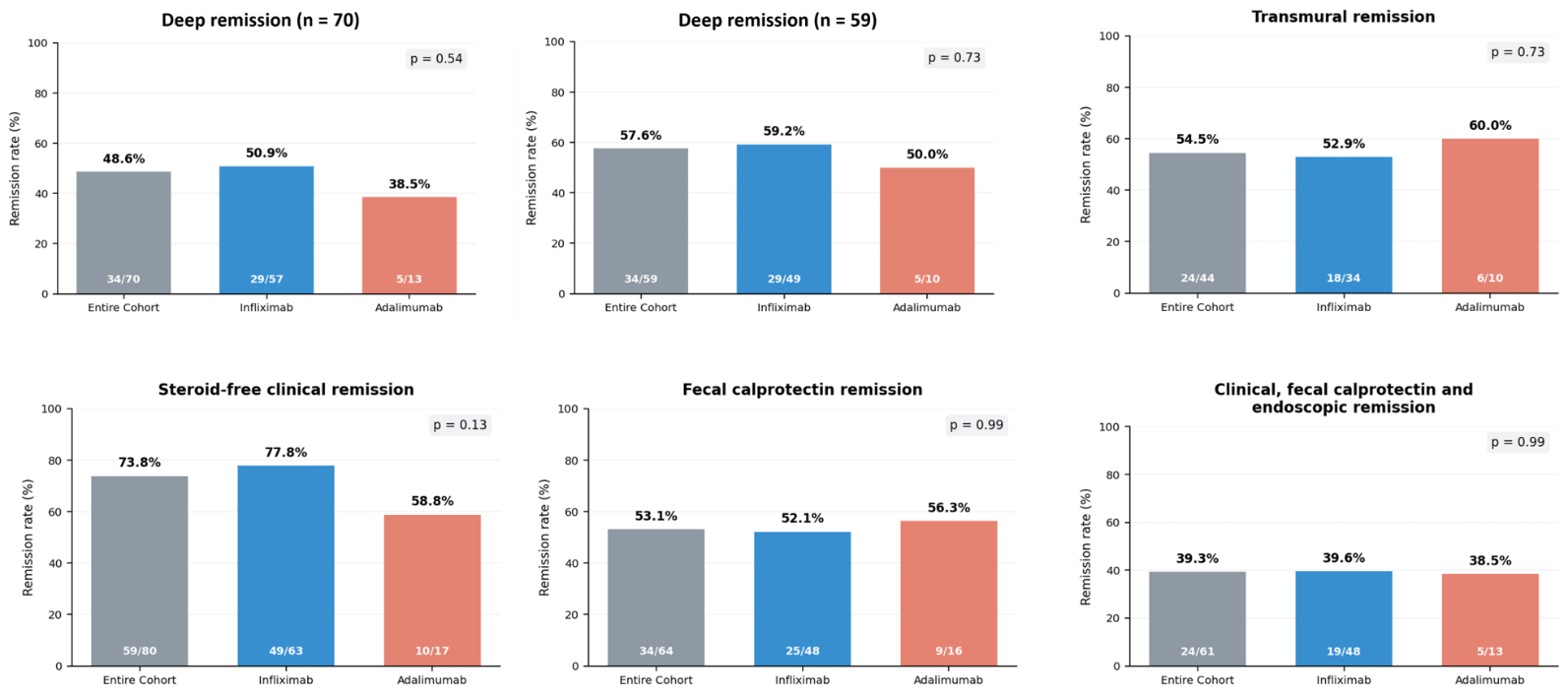
**Supplemental Figure 2. Month12 treatment outcomes**. Deep remission (n=70) included participants who underwent follow-up colonoscopy or discontinued anti-TNF therapy or underwent CD-related surgery before month12. Deep remission (n=59) included only participants who underwent follow-up colonoscopy, excluding those without colonoscopy or discontinued anti-TNF therapy or underwent surgery.

**Supplemental Figure 3:** Mean-variance plots of normalized protein measurements at baseline (Panel A), month3 (Panel B), and month 12 (Panel C).

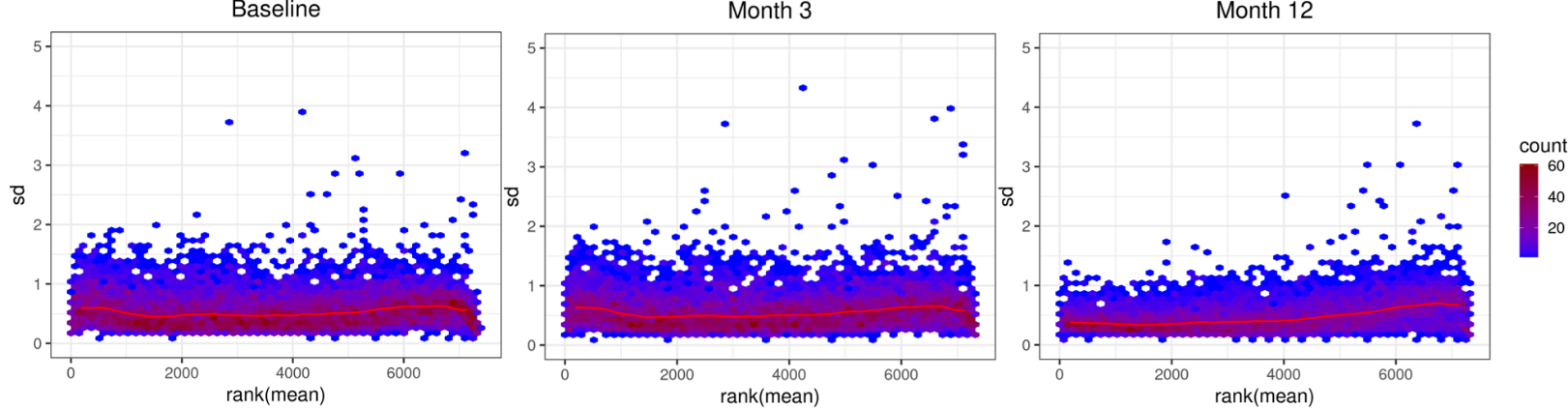

**Supplemental Table 1.** Baseline demographics, disease characteristics, and laboratory results for all subjects and those who achieved deep remission and those who did not achieve deep remission.

| **Characteristic** | **All patients**  **(n=80)** | **Deep Remission (n=34)** | **No-Deep Remission (n=36)** | **p-value** |
| --- | --- | --- | --- | --- |
| Age at start of therapy, years (mean, SD) | 13.7 (3.5) | 13.4 (3.2) | 13.5 (3.7) | 0.90 |
| Age at diagnosis, years (mean, SD) | 13.4 (3.6) | 13 (3.3) | 13.4 (3.8) | 0.69 |
| Gender, female | 34 (42.5%) | 15 (44.1%) | 13 (36.1%) | 0.63 |
| Race, white | 72 (90%) | 31 (91.2%) | 32 (88.9%) | 0.99 |
| Ethnicity, non-Hispanic | 78 (97.5%) | 34 (100%) | 34 (94.4%) | 0.49 |
| Body mass index, kg/m^2^ | 17 (15-21) | 17.5 (15.5-21.8) | 17.5 (15-20.8) | 0.96 |
| Methotrexate combination from start | 8 (10%) | 5 (14.7%) | 2 (5.6%) | 0.25 |
| Baseline corticosteroid exposure | 37 (46.3%) | 15 (44.1%) | 17 (47.2%) | 0.82 |
| Prednisone : Budesonide | 19 : 18 | 7 : 8 | 10 : 7 | - |
| **Type of Anti-TNF** |  |  |  |  |
| Infliximab : Adalimumab | 63 : 17 | 29 : 5 | 28 : 8 | 0.54 |
| Infliximab mg/kg starting dose | 6.5 (5.2-9.2) | 5.9 (5.2-8.7) | 7.4 (5.4-9.3) | 0.21 |
| Adalimumab mg/kg starting dose (mean, SD) | 3 (0.68) | 3.7 (0.42) | 2.8 (0.63) | **0.011** |
| Time to anti-TNF, days | 31 (16-48) | 35 (22-53) | 28 (14-37) | **0.037** |
| Less than 90 days form diagnosis | 72 (90%) | 30 (88.2%) | 33 (91.7%) | 0.71 |
| **Crohn’s Location** |  |  |  |  |
| L1 : L2 : L3 | 26 : 10 : 44 | 10 : 4 : 20 | 12 : 5 : 19 | - |
| Perianal phenotype | 17 (21.3%) | 5 (14.7%) | 10 (27.8%) | 0.25 |
| **Crohn’s behavioral** |  |  |  |  |
| B1 : B2 : B3 : B4 | 66 : 11 : 3 : 0 | 31 : 2 : 1 : 0 | 28 : 8 : 0 : 0 | - |
| Weighted Pediatric CD activity index | 33 (18-50) | 24 (10-45) | 43 (25-53) | **0.037** |
| Albumin, g/dL | 3.6 (3.3-4) | 3.6 (3.1-4.1) | 3.5 (3.3-4) | 0.66 |
| Erythrocyte sedimentation rate, mm/h | 28 (12-40) | 18 (11-29) | 33 (11-65) | 0.054 |
| C-reactive protein, mg/dL | 1.4 (0.58-2.6) | 1.2 (0.4-2.4) | 1.6 (0.96-2.7) | 0.18 |
| Neutrophil CD64 ratio | 6 (4-9.4) | 5.9 (3.9-9.4) | 6 (3.9-9.7) | 0.98 |
| Soluble CD64, ng/mL (mean, SD) | 27.1 (8.7) | 27.4 (9.3) | 26.9 (8.2) | 0.82 |
| Fecal calprotectin, µg/g | 2102 (878-3694) | 1873 (761-3112) | 2584 (862-4126) | 0.08 |

Data shown as number (%) or median (25-75 interquartile range) unless otherwise specified as mean (SD, standard deviation).

P-value represents the comparison between deep remission and no-deep remission

**Supplemental Table 2. Early (Month3) and Late (Month12) Outcomes by Anti-TNF therapy.**

| **Outcome** | **Entire Cohort** | **Infliximab** | **Adalimumab** | **p-value** |
| --- | --- | --- | --- | --- |
| **Month 3 Outcomes** |  |  |  |  |
| Primary clinical nonresponse | 9/80 (11.3%) | 6/63 (9.5%) | 3/17 (17.7%) | 0.39 |
| Primary biochemical nonresponse | 13/72 (18.1%) | 13/58 (22.4%) | 0/14 (0%) | 0.06 |
| Steroid-free clinical remission | 49/80 (61.3%) | 39/63 (61.9%) | 10/17 (58.8%) | 0.99 |
| Fecal calprotectin remission | 29/63 (46%) | 24/49 (49%) | 5/14 (35.7%) | 0.55 |
| Biochemical remission | 42/80 (52.5%) | 34/63 (54%) | 8/17 (47.1) | 0.79 |
| Adequate anti-TNF exposure | 42/70 (60%) | 30/54 (55.6%) | 12/16 (75%) | 0.25 |
| *Exploratory* |  |  |  |  |
| *Fecal calprotectin healing (<150 µg/g)* | 23/63 (36.5%) | 18/49 (36.7%) | 5/14 (35.7%) | 0.99 |
| *Ideal anti-TNF exposure* | 39/70 (55.7%) | 32/54 (59.3%) | 7/16 (43.8%) | 0.39 |
| **Month12 Outcomes** |  |  |  |  |
| Deep remission (n=70)^1^ | 34/70 (48.6%) | 29/57 (50.9%) | 5/13 (38.5%) | 0.54 |
| Deep remission (n=59)^2^ | 34/59 (57.6%) | 29/49 (59.2%) | 5/10 (50%) | 0.73 |
| Steroid-free clinical remission | 59/80 (73.8%) | 49/63 (77.8%) | 10/17 (58.8%) | 0.13 |
| Endoscopic remission (n=59)^3^ | 35/59 (59.3%) | 30/49 (61.2%) | 5/10 (50%) | 0.73 |
| Endoscopic remission (n=70)^4^ | 35/70 (50%) | 30/57 (52.6%) | 5/13 (38.5%) | 0.54 |
| Fecal calprotectin remission | 34/64 (53.1%) | 25/48 (52.1%) | 9/16 (56.3%) | 0.99 |
| Fecal calprotectin and endoscopic remission | 25/60 (41.7%) | 20/47 (42.6%) | 5/13 (38.5%) | 0.99 |
| Clinical, fecal calprotectin and endoscopic remission | 24/61 (39.3%) | 19/48 (39.6%) | 5/13 (38.5%) | 0.99 |
| Transmural remission | 24/44 (54.5%) | 18/34 (52.9%) | 6/10 (60%) | 0.73 |
| Adequate anti-TNF exposure | 45/64 (70.3%) | 36/50 (72%) | 9/14 (64.3%) | 0.74 |
| *Exploratory* |  |  |  |  |
| *Fecal calprotectin healing* | 28/64 (43.8%) | 20/48 (41.7%) | 8/16 (50%) | 0.58 |
| *Transmural healing* | 22/49 (44.9%) | 18/38 (47.4%) | 4/11 (36.4%) | 0.73 |
| *Ideal anti-TNF exposure* | 43/64 (67.2%) | 36/50 (72%) | 7/14 (50%) | 0.20 |

Definition of each outcome is noted in the Methods. ^1^Participants who had a follow-up colonoscopy (SES-CD<3) and a wPCDAI<12.5 or either discontinued anti-TNF therapy or underwent CD-related surgery before month12. ^2^Participants who had a follow-up colonoscopy (SES-CD<3) and a wPCDAI<12.5 and remained on anti-TNF therapy without surgery at month12. ^3^Inclusive of participants who had a follow-up colonoscopy (SES-CD<3) and remained on anti-TNF therapy without surgery at month12. ^4^Inclusive of participants who had a follow-up colonoscopy (SES-CD<3), stopped anti-TNF therapy prior to month12 or had surgery before month12.
